# Association between Postoperative Anastomotic Leakage and Dynamic Changes of SII Score in Esophageal Cancer Patients With Neoadjuvant Chemoradiotherapy : A two-center study

**DOI:** 10.1101/2025.11.13.25340188

**Authors:** Mingchao Wei, Hai Zhang, MaoXiu Yuan, ZhenYang Zhang, Jiangbo Lin

## Abstract

**Objective:** To investigate the correlation between postoperative anastomotic leakage (AOL) and dynamic changes in systemic immune inflammatory index (SII) scores in esophageal cancer patients undergoing neoadjuvant chemoradiotherapy (NAC-CA) after surgery.

**Methods:** A retrospective analysis was conducted on 247 esophageal cancer patients who underwent NAC-CA surgery at Fujian Medical University Union Hospital and Yantai Affiliated Hospital of Binzhou Medical University (two centers) from January 2021 to December 2023. Patients were classified into two groups based on postoperative AOL occurrence: leakage group (38 cases) and non-leakage group (209 cases). The study compared general demographics and dynamic SII scores (preoperative, postoperative day 1, day 3, day 7) between groups, and performed multivariate logistic regression analysis to identify independent risk factors for AOL development.

**Results:** The leakage group showed significantly higher rates of age, hypertension prevalence, and dynamic SII scores (especially postoperative day 3) compared to the non-leakage group (P<0.05). The SII score of the leakage group peaked on postoperative day 3 (1897.0±592.9), which was significantly higher than that of the non-leakage group (1144.5±316.7) on the same day. Multivariate logistic regression analysis revealed that age (OR=1.05,95% CI=1.02-1.08, P=0.00), hypertension (OR=2.49,95%CI=1.09-5.67, P=0.03), and postoperative day 3 SII score (OR=1.003,95%CI=1.003-1.004, P=0.00) were independent risk factors for AOL.

**Conclusion:** Dynamic changes of SII scores in esophageal cancer patients undergoing NAC-CA are closely associated with AOL occurrence, and the postoperative day 3 SII score is the most valuable predictor for assessing postoperative leakage risk.

## 1. Introduction

Data from the National Cancer Center indicates that esophageal cancer ranks sixth in incidence and fifth in mortality among malignant tumors detected in China, with higher prevalence rates observed in males than females [1]. While surgical resection can improve patients’ quality of life and prolong survival time to some extent, relying solely on surgery carries a relatively high failure rate [2]. According to the CROSS study, neoadjuvant chemoradiotherapy has become the standard treatment for locally advanced resectable esophageal cancer patients or those with gastroesophageal junction cancer. The long-term survival results from the 2021 CROSS study revealed that patients receiving neoadjuvant concurrent chemoradiotherapy achieved a 10-year survival rate of 38% [3].

The systemic immune inflammation index (SII) is calculated based on the counts of platelets, neutrophils, and lymphocytes. It can provide a comprehensive assessment of the immune status and inflammatory balance of the body. Studies have shown that SII is related to the prognosis of various cancers and postoperative complications. However, for patients with esophageal cancer undergoing neoadjuvant chemoradiotherapy, there are relatively few studies on the relationship between dynamic changes of SII and anastomotic leakage. This article aims to explore the dynamic correlation between AL and SII scores (preoperative, postoperative day 1, day 3, day 7) in patients with esophageal cancer after neoadjuvant chemoradiotherapy, in order to provide clinical guidance for the evaluation and prevention of AL.

## 2. Patients and methods

### 2.1 Patients

Retrospective collection of clinical data from esophageal cancer patients undergoing neoadjuvant chemoradiotherapy at Fujian Medical University Union Hospital Thoracic Surgery Department and Yantai Affiliated Hospital of Binzhou Medical University Thoracic Surgery Department (two centers) from January 1, 2021, to December 31, 2023. The study was conducted in accordance with the Declaration of Helsinki (as revised in 2013). Verbal informed consent was obtained from all participants via telephone.Each consent process was documented by two independent researchers (including call date, communicated information, and participant’s agreement) and archived. All identifiable information was anonymized prior to data analysis to protect participant privacy. The study was approved by the Institutional Review Board of Fujian Medical University Union Hospital (ethical No. 2024KY037) and Yantai Affiliated Hospital of Binzhou Medical University (ethical No. 20251104160).

Inclusion criteria: (1) Pathologically confirmed esophageal squamous cell carcinoma; (2) Preoperative neoadjuvant chemoradiotherapy (including immune checkpoint inhibitors combined with chemotherapy); (3) Esophageal cancer radical resection with gastrointestinal reconstruction; (4) Complete clinical and follow-up data, including preoperative and postoperative serial SII test results (day 1, day 3, day 7 after surgery). Exclusion criteria: (1) Concurrent malignancies; (2) Preoperative severe infections (e.g., pneumonia, sepsis), autoimmune diseases; (3) Preoperative radiotherapy, chemotherapy, or other anti-tumor treatments; (4) Death within 30 days postoperative; (5) Postoperative pulmonary complications (e.g., atelectasis, suspected pneumonia) where clinical data (e.g., missing dynamic imaging follow-ups, pathogen detection results, or inflammatory marker monitoring) were insufficient for guideline-compliant differential diagnosis.

The differentiation between postoperative pneumonia and atelectasis remains a common clinical challenge in esophageal cancer perioperative care. Given their overlapping symptoms (e.g., low-grade fever, mild cough) and imaging features (such as localized pulmonary infiltrates), this study did not directly include postoperative pneumonia in exclusion criteria (to avoid missing real cases with concurrent anastomotic leakage and pulmonary complications, thereby reducing selection bias). Instead, we strictly required "complete documentation of pulmonary status" and referenced the differentiation criteria from the "Chinese Expert Consensus on Perioperative Pulmonary Complications in Esophageal Cancer (2023 Edition)". The pneumonia/atelectasis diagnosis was established through: ① fever>38.0 ℃ or hypothermia <36.0℃ with peripheral white blood cell count>10×10⁹/L or <4×10⁹/L; ② cough, sputum production, or dyspnea; ③ chest CT showing new inflammatory infiltrates; ④ positive pathogen testing (sputum culture, blood culture) or clinical evidence of infection. Atelectasis was diagnosed based on imaging findings of reduced lung air content and tissue collapse, without abnormal infection-related indicators. This approach ensured clear categorization of postoperative pulmonary lesions, allowing subsequent analysis to control for potential confounding effects of postoperative pneumonia on SII scoring and anastomotic leakage correlation.

Finally, 247 patients were included, including 186 males (75.3%) and 61 females (24.7%), with an average age of 42-78 years (61.2±8.5). Among the 247 patients, 21 cases (8.5%) were diagnosed as pneumonia after surgery, 43 cases (17.4%) were simple atelectasis, and 183 cases (74.1%) had no obvious lung lesions.

### 2.2 Methodology

#### 2.2.1 Data collection

Patient data were extracted from the hospital electronic medical record system. In this retrospective study, all the data were de-identified by the data provider (e.g., the hospital’s medical record department, the research repository) prior to access. During and after data collection, the research team had no access to any information that could identify individual participants, including but not limited to names, ID numbers, hospital record numbers, phone numbers, and residential addresses.

Data including:

General information: gender, age, height, weight, BMI, smoking history, drinking history, previous medical history (hypertension, diabetes, coronary heart disease, etc.);

Treatment related: neoadjuvant chemotherapy regimen, cycle, surgical method and time;

Laboratory indicators: preoperative SII score and postoperative SII scores at day 1, day 3, day 7 (SII= platelet count × neutrophil count/lymphocyte count), carcinoembryonic antigen (CEA), squamous epithelial cell carcinoma antigen (SCC); Postoperative complications: focus on recording the occurrence and classification of anastomotic leakage, and the time of leakage occurrence (to correlate with SII dynamic changes).

#### 2.2.2 Diagnosis and classification of anastomotic leakage

Diagnostic criteria for anastomotic leakage: (1) fever, chest pain, dyspnea and other symptoms after surgery; (2) imaging examination (angiography, CT) indicates fluid accumulation or external leakage of contrast agent; (3) endoscopic examination shows anastomotic defect. Grading criteria [4]:

Grade I: asymptomatic, only imaging suggests leakage;

Grade Ⅱ: mild symptoms (fever, elevated white blood cells), localized leakage;

Grade Ⅲ : obvious symptoms (septicemia), endoscopy showed anastomosis defect;

Grade Ⅳ: Esophageal replacement organ necrosis.

#### 2.2.3 Statistical methods

The data were analyzed using SPSS 26.0 software. Quantitative data were presented as mean ± standard deviation (x ± s), with inter-group comparisons conducted using the independent samples t-test. Repeated-measures ANOVA was used to compare the dynamic changes of SII scores at different time points (preoperative, postoperative day 1, day 3, day 7) within and between groups. Categorical data were expressed as frequency (%) and compared using χ ² test. Multivariate logistic regression was employed to analyze independent risk factors for anastomotic leakage. ROC curve analysis was used to evaluate the predictive value of SII scores at different time points for anastomotic leakage. P values <0.05 indicated statistically significant differences.

## 3. Results

### 3.1 General information and grouping of patients

Among the 247 patients, anastomotic leakage occurred in 38 cases (leak group) and no anastomotic leakage occurred in 209 cases (no leak group). The general data comparison between the two groups is shown in Table 1.

**Table 1.** Comparison of general data between two groups.

| Variables | Leakage group<br>(n=38) | No leakage group (n=209) | Statistic | P value |
| --- | --- | --- | --- | --- |
| Gender (male/female, n) | 30/8 | 156/53 | $\chi^2=0.89$ | 0.34 |
| Age (years, $x \pm s$) | $65.4 \pm 7.2$ | $59.8 \pm 8.6$ | $t=4.21$ | 0.00 |
| Height (cm, $\bar{x} \pm s$) | 165.2±6.8 | 166.5±7.1 | t=1.01 | 0.41 |
| Weight (kg, $\bar{x} \pm s$) | 58.6±8.3 | 61.2±9.1 | t=1.64 | 0.10 |
| Smoking (Yes/No, n) | 28/10 | 152/57 | $\chi^2=0.00$ | 1.00 |
| Drinking (Yes/No, n) | 26/12 | 145/64 | $\chi^2=0.89$ | 0.34 |
| Hypertension (Yes/No, n) | 18/20 | 56/153 | $\chi^2=4.76$ | 0.02 |
| Diabetes (Yes/No, n) | 6/32 | 35/174 | $\chi^2=0.24$ | 0.62 |

As shown in Table 1, the age of patients in the leakage group was significantly higher than that in the non-leakage group (P=0.00), and the prevalence of hypertension was significantly higher than that in the non-leakage group (P=0.02); other indicators showed no statistical significance between the two groups (P>0.05).

### 3.2 Dynamic comparison of SII scores between the two groups

The postoperative SII score of the leakage group (1897.0 ± 592.9) was significantly higher than that of the non-leakage group (1144.5 ± 316.7), and the difference was statistically significant (t=12.27, P=0.00).

### 3.3 Comparison of SII scores in patients with different grades of anastomotic leakage

The dynamic changes of SII scores in the two groups at different time points are shown in Table 2 and Figure 1. Within the leakage group, the SII score showed a trend of increasing first and then decreasing, peaking on postoperative day 3 (1897.0 ±592.9); while the non-leakage group showed a mild increase on postoperative day 1 and then gradually decreased, with the peak value on day 1 (1210.3 ± 356.2). Repeated-measures ANOVA showed that the main effect of group was significant (F=102.36, P<0.001), the main effect of time was significant (F=45.72, P<0.001), and the interaction effect between group and time was significant (F=28.91, P<0.001). Post hoc analysis showed that the SII scores of the leakage group were significantly higher than those of the non-leakage group at postoperative day 1, day 3, and day 7 (all P<0.05), and the difference was most significant on day 3 (t=12.27, P=0.00).

**Figure 1:**
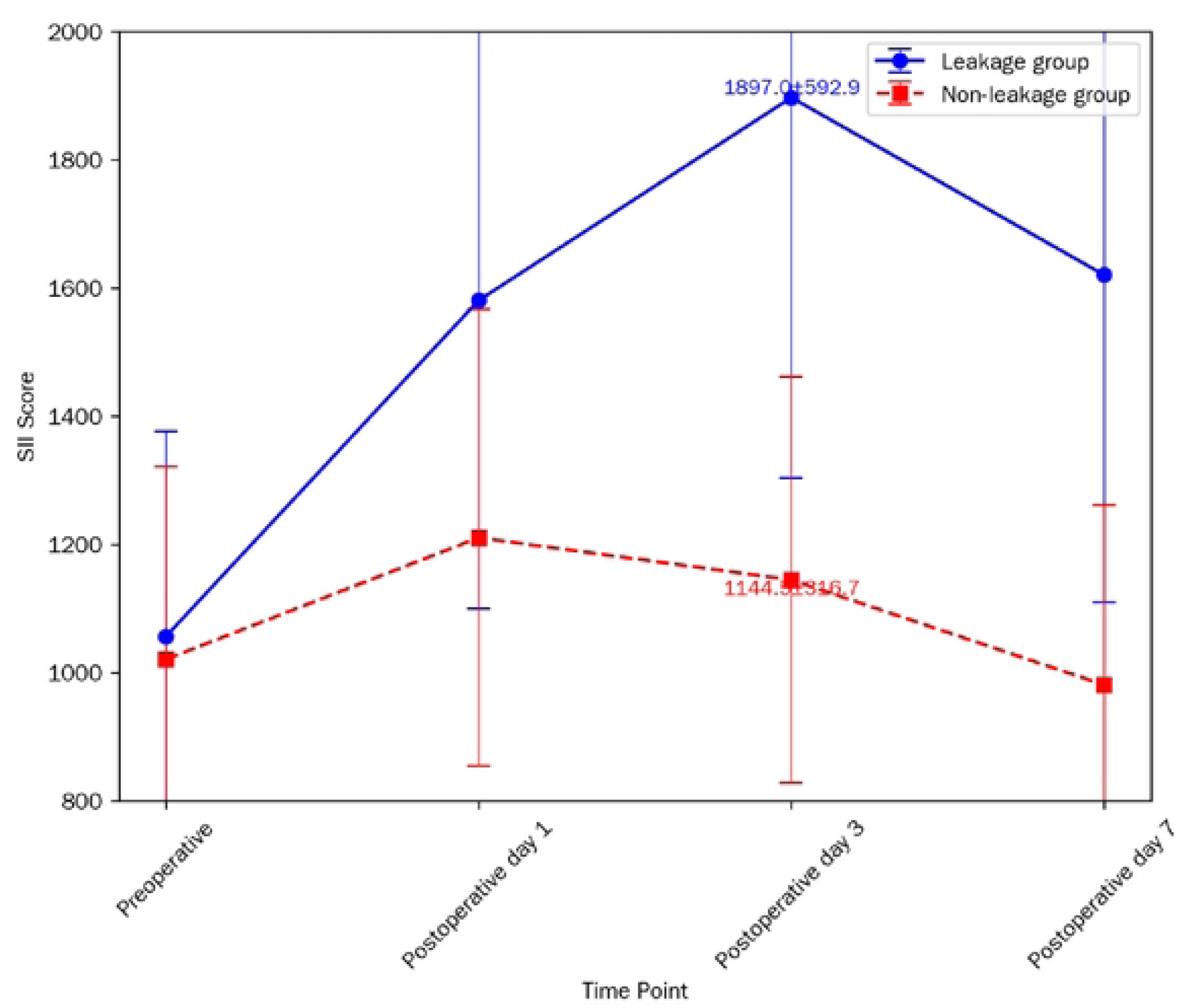
Dynamic changes of SII scores in two groups at different time points. Note: The horizontal axis represents different time points (preoperative, postoperative day 1, day 3, day 7), and the vertical axis represents SII score. The error bar represents the standard deviation. The leakage group shows a significant peak on postoperative day 3, which is significantly higher than the non-leakage group. Postoperative day 3 is the key time point for SII score to predict anastomotic leakage.

**Table 2.** Dynamic comparison of SII scores between two groups (x±s)

| Group | n | Preoperative | Postoperative day<br>1 | Postoperative day<br>3 | Postoperative day<br>7 |
| --- | --- | --- | --- | --- | --- |
| Leakage group | 38 | 1056.2±<br>320.4 | 1580.5±480.7 | 1897.0±592.9 | 1620.3±510.6 |
| Non-leakage<br>group | 209 | 1020.5±<br>300.8 | 1210.3±356.2 | 1144.5±316.7 | 980.6±280.3 |
| t value | - | 0.68 | 5.92 | 12.27 | 8.35 |
| P value | - | 0.50 | 0.00 | 0.00 | 0.00 |

### 3.3 Comparison of SII scores in patients with different grades of anastomotic leakage

Among 38 patients with anastomotic leakage, there were 8 cases of grade I, 15 cases of grade II, 12 cases of grade III and 3 cases of grade IV. The SII scores at postoperative day 3 (the peak time point) among patients with different grades are shown in Table 3. With the increase of anastomotic leakage grade, the SII score at postoperative day 3 showed an upward trend: grade I (1420.5 ± 380.2), grade II (1810.3±490.5), grade III (2150.7±560.3), grade IV (2380.5±610.9). However, the difference was not statistically significant (F=0.68, P=0.57), which may be related to the small number of cases in grade IV.

**Table 3.**
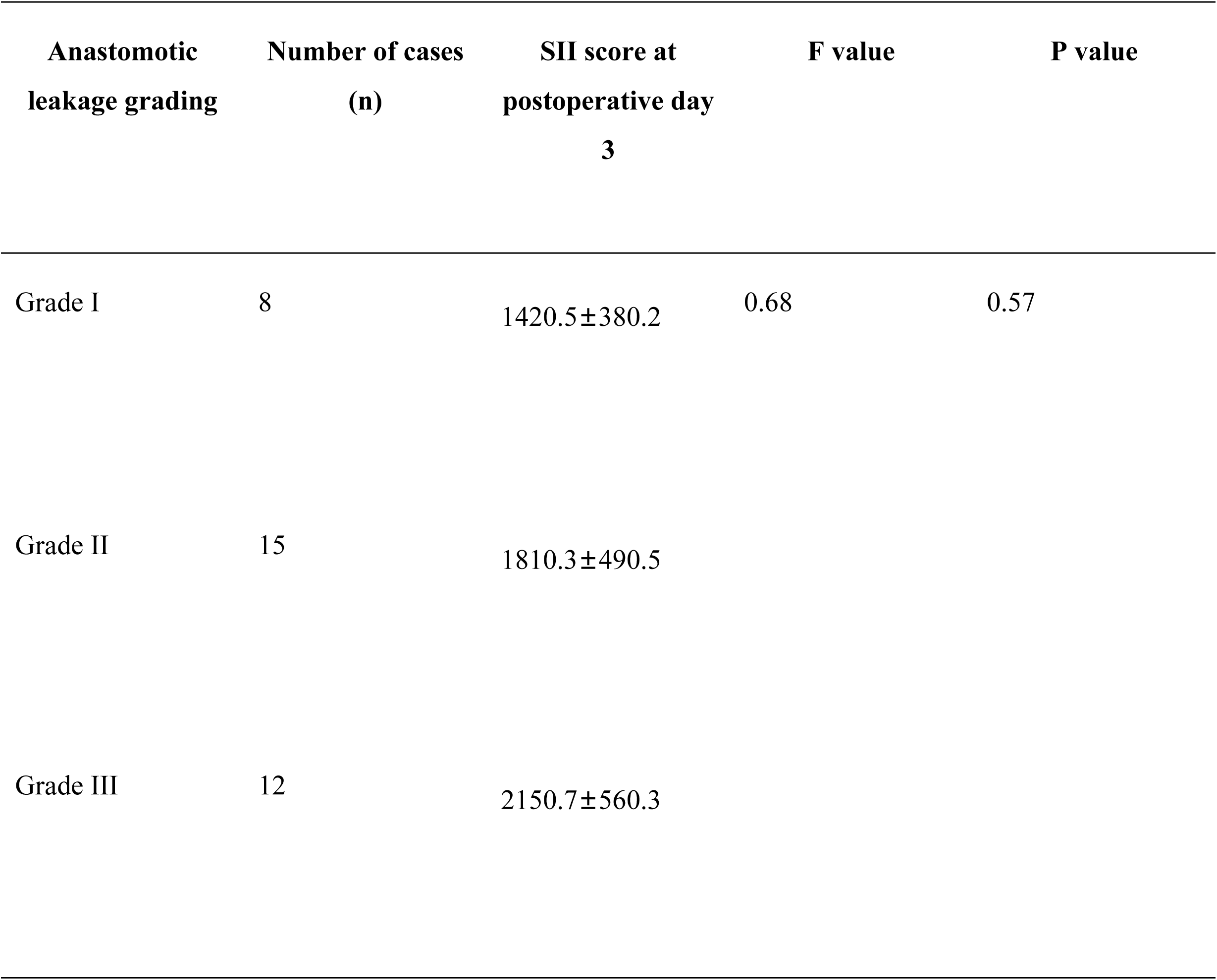
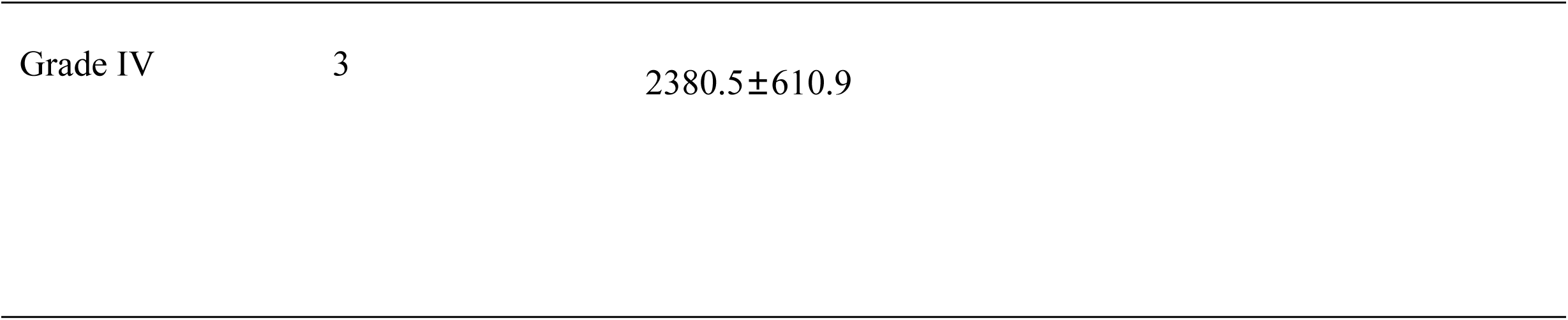
Comparison of SII scores at postoperative day 3 in patients with different grades of anastomotic leakage (x ±s)

| Anastomotic leakage grading | Number of cases (n) | SII score at postoperative day 3 | F value | P value |
| --- | --- | --- | --- | --- |
| Grade I | 8 | $1420.5 \pm 380.2$ | 0.68 | 0.57 |
| Grade II | 15 | $1810.3 \pm 490.5$ | | |
| Grade III | 12 | $2150.7 \pm 560.3$ | | |
| Grade IV | 3 | 2380.5±610.9 |  |  |

### 3.4 Multivariate Logistic regression analysis of the occurrence of anastomotic leakage

The variables with P <0.1 in the univariate analysis (age, hypertension, SII scores at postoperative day 1, day 3, day 7) were included in the multivariate Logistic regression model. The results showed that age, hypertension and SII score at postoperative day 3 were independent risk factors for postoperative anastomotic leakage in esophageal cancer patients after neoadjuvant chemoradiotherapy (P <0.05), as shown in Table 4.

**Table 4.** Multivariate logistic regression analysis of the occurrence of anastomotic leakage.

| Variable | Coefficient | Standard error | z value | P value | OR value | 95%CI |
| --- | --- | --- | --- | --- | --- | --- |
| Constant term | -6.15 | 1.19 | -5.17 | 0.00 | - | - |
| Age | 0.05 | 0.02 | 2.93 | 0.00 | 1.05 | 1.02-1.08 |
| Hypertension | 0.91 | 0.42 | 2.17 | 0.03 | 2.49 | 1.09-5.67 |
| SII score at postoperative day 3 | 0.003 | 0.00 | 11.29 | 0.00 | 1.003 | 1.003-1.004 |

### 3.5 Predictive value of SII scores at different time points for postoperative anastomotic leakage

ROC curve analysis was performed to compare the predictive value of SII scores at different time points for postoperative anastomotic leakage. The results showed that the AUC of SII score at postoperative day 3 was 0.876 (95% CI=0.815-0.937, P=0.00), which was significantly higher than that at preoperative (AUC=0.523, P=0.68), postoperative day 1 (AUC=0.752, P=0.00) and day 7 (AUC=0.801, P=0.00). According to the maximum Youden index method, the optimal cutoff value of SII score at postoperative day 3 for predicting anastomotic leakage was 1526.3, with a sensitivity of 81.6% (66.4%-91.5%) and a specificity of 83.3% (78.0%-87.7%), as shown in Table 5 and Figure 2.

**Figure 2:**
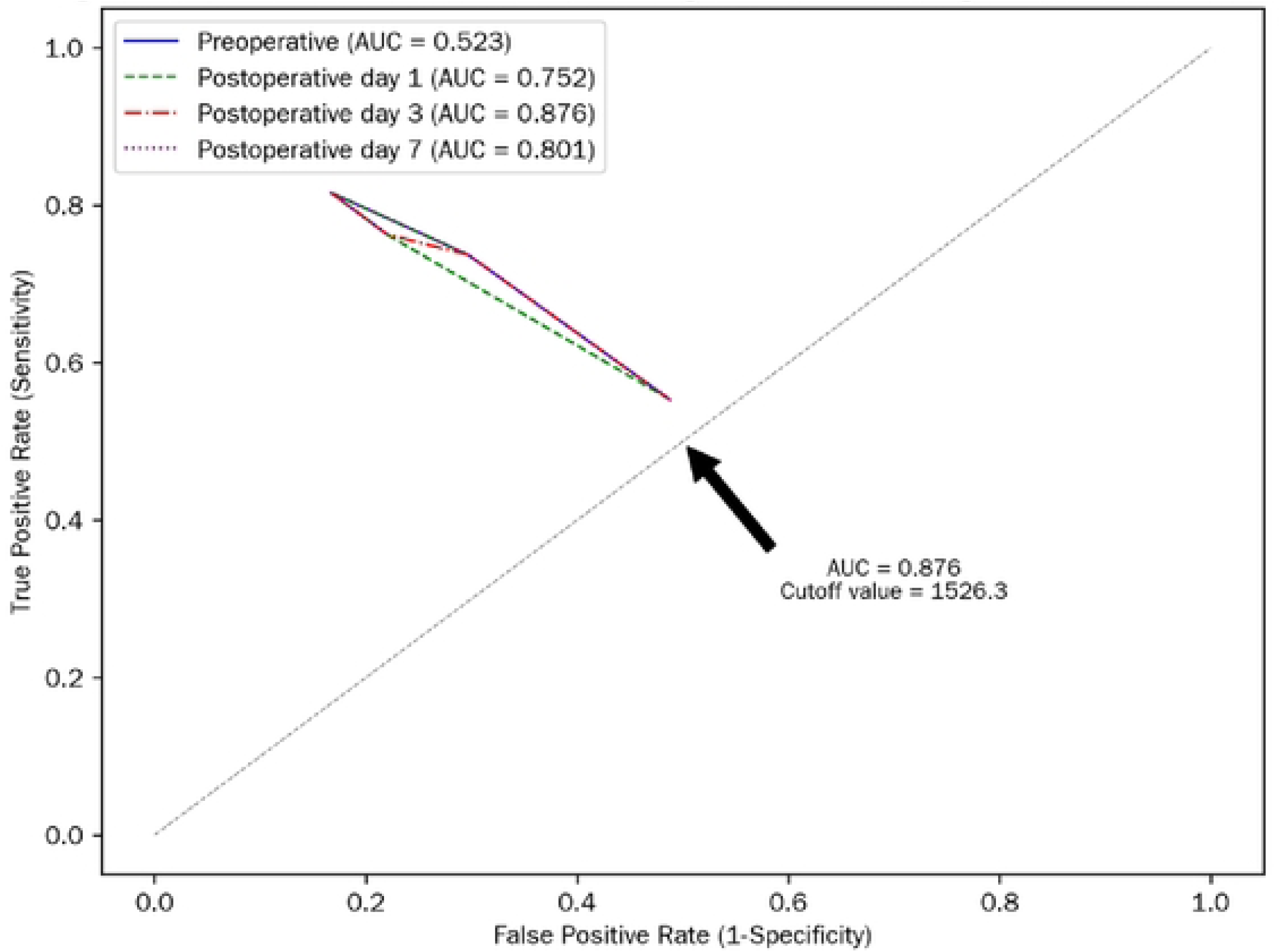
ROC curves of SII scores at different time points for predicting postoperative anastomotic leakage. Note: The horizontal axis represents 1-specificity (false positive rate), and the vertical axis represents sensitivity (true positive rate). The curve of SII score at postoperative day 3 is the closest to the upper left corner, with the largest AUC (0.876), indicating the best predictive performance. The arrow marks the optimal cutoff value (SII=1526.3) at postoperative day 3.

**Table 5.** ROC curve performance indexes of SII scores at different time points for predicting postoperative anastomotic leakage.

| Time point | AUC<br>(95%CI) | P<br>value | Optimal<br>cutoff<br>value | Sensitivity<br>(95%CI) | Specificity<br>(95%CI) | Youden<br>index |
| --- | --- | --- | --- | --- | --- | --- |
| Preoperative | 0.523<br>(0.431-0.615) | 0.68 | 1080.2 | 55.3%<br>(38.6%-71.2%) | 51.2%<br>(44.3%-58.1%) | 0.065 |
| Postoperative<br>day 1 | 0.752<br>(0.678-0.826) | 0.00 | 1350.5 | 73.7%<br>(57.9%-85.9%) | 70.3%<br>(63.8%-76.3%) | 0.440 |
| Postoperative<br>day 3 | 0.876<br>(0.815-0.937) | 0.00 | 1526.3 | 81.6%<br>(66.4%-91.5%) | 83.3%<br>(78.0%-87.7%) | 0.649 |
| Postoperative<br>day 7 | 0.801<br>(0.732-0.870) | 0.00 | 1380.7 | 76.3%<br>(60.8%-87.9%) | 78.0%<br>(71.7%-83.6%) | 0.543 |

Conclusion: Postoperative day 3 is the most valuable time point for SII score to predict anastomotic leakage, with high sensitivity and specificity.

## 4. Discussion

### 4.1 The correlation mechanism and clinical significance between SII score and anastomotic leakage

The results of this study demonstrate that esophageal cancer patients who developed anastomotic leakage after neoadjuvant chemoradiotherapy showed significantly higher dynamic SII scores (preoperative, postoperative day 1, day 3, day 7) compared to those without leakage, with the most significant difference on postoperative day 3 (leakage group: 1897.0 ± 592.9 vs. non-leakage group: 1144.5 ± 316.7, P=0.00). Multivariate logistic regression analysis confirmed that postoperative day 3 SII score is an independent risk factor for anastomotic leakage (OR=1.003,95%CI=1.003-1.004, P=0.00), and ROC curve analysis showed its AUC reached 0.876 (95%CI=0.815-0.937), significantly higher than other time points. These findings reveal that dynamic changes of SII score, as a comprehensive indicator reflecting immune-inflammatory status, are closely associated with postoperative anastomotic healing in esophageal cancer patients, and postoperative day 3 is the optimal predictive time point.

The SII score integrates platelet, neutrophil, and lymphocyte counts to assess the body’s inflammatory response and immune balance. After esophageal cancer surgery, anastomotic healing involves inflammation, cell proliferation, and collagen synthesis. Excessive neutrophil activation causes local tissue damage and edema, abnormal platelet elevation leads to thrombosis and impaired blood supply, and reduced lymphocytes weaken immune defense against infection—all contributing to an imbalanced immune-inflammatory state and elevated SII score that hinders anastomotic healing.

From a clinical perspective, the SII score is a convenient, reusable assessment tool for early evaluation of anastomotic leakage risk in this patient population. Based on the optimal cutoff value of 1526.3 for postoperative day 3 SII score (sensitivity 81.6%, specificity 83.3%), clinicians can identify high-risk patients early. Targeted interventions include suppressing excessive inflammation with anti-inflammatory drugs, enhancing immune function through nutritional support, and closely monitoring infection indicators for timely antibacterial treatment, which may reduce leakage incidence or alleviate severity.

### 4.2 Influence of age on anastomotic leakage and countermeasures

This study found that age is an independent risk factor for postoperative anastomotic leakage in esophageal cancer patients receiving neoadjuvant chemoradiotherapy (OR=1.05,95% CI=1.02-1.08, P=0.00), with the leakage group’s average age (65.4±7.2 years) significantly higher than the non-leakage group (59.8± 8.6 years, P=0.00). This indicates the risk of anastomotic leakage increases by 5% for every additional year of age, aligning with previous research and highlighting age’s significant impact on postoperative healing.

Aging leads to gradual decline in systemic function, reduced tolerance to surgical trauma, and impaired tissue repair ability. Elderly patients often have vascular hardening and decreased elasticity, resulting in insufficient anastomotic blood supply that disrupts nutrient delivery and metabolic waste clearance, delaying wound healing. Their weakened immune systems—characterized by reduced T lymphocyte proliferation and cytokine secretion—lower infection resistance, increasing anastomotic infection risk and subsequent leakage. Additionally, elderly patients are more likely to have comorbidities such as hypertension, further elevating complication risks.

For elderly patients, clinical management should adopt personalized strategies. Preoperatively, conduct comprehensive physical assessments to improve nutritional status (high-protein, high-calorie diet) and immune function, with immunostimulants administered if necessary. Intraoperatively, use meticulous surgical techniques to protect anastomotic blood supply. Postoperatively, strengthen monitoring of anastomotic healing to detect early complications like infection or malnutrition for timely intervention.

### 4.3 The association and management of hypertension and anastomotic leakage

This study confirms that hypertension is an independent risk factor for postoperative anastomotic leakage in esophageal cancer patients receiving neoadjuvant chemoradiotherapy (OR=2.49,95% CI=1.09-5.67, P=0.03). The prevalence of hypertension in the leakage group (47.4%, 18/38) was significantly higher than in the non-leakage group (26.8%, 56/209, P=0.02), indicating hypertensive patients face 2.49 times higher leakage risk. Hypertension’s adverse effects on anastomotic healing are primarily linked to vascular lesions.

Chronic hypertension causes systemic arteriosclerosis and endothelial damage, reducing vascular wall elasticity and lumen diameter, which impairs tissue blood perfusion. The anastomotic site’s inherently fragile blood supply after esophageal cancer surgery is further compromised by hypertension-related vascular changes, leading to local ischemia and hypoxia that disrupts cell metabolism and collagen synthesis, delaying healing and increasing leakage risk. Hypertension may also disrupt coagulation function, elevating anastomotic thrombosis risk and exacerbating ischemia.

For hypertensive esophageal cancer patients, preoperative blood pressure control below 140/90 mmHg is recommended. Clinicians select appropriate antihypertensive drugs (calcium channel blockers, angiotensin-converting enzyme inhibitors, etc.) and monitor blood pressure fluctuations closely. Intraoperatively, maintain stable blood pressure to avoid anastomotic hypoperfusion (from hypotension) or increased tension (from hypertension). Postoperatively, continue strengthening blood pressure control through regular monitoring and timely antihypertensive plan adjustments to maintain stability and create a favorable environment for anastomotic healing.

### 4.4 Potential effects of neoadjuvant chemoradiotherapy on SII scores and anastomotic leakage risk

Existing studies have shown that neoadjuvant chemoradiotherapy (including immune checkpoint inhibitors combined with chemotherapy) significantly improves the major pathological response (MPR) and pathological complete response (pCR) rates of locally advanced esophageal cancer compared to neoadjuvant chemotherapy alone, becoming the standard treatment for resectable cases [5-6]. This treatment effectively inhibits tumor progression and reduces tumor size but may alter the body’s immune-inflammatory status, indirectly affecting SII scores and postoperative anastomotic leakage risk.

Immune checkpoint inhibitors (e.g., PD-1/PD-L1 blockers) activate T lymphocytes by reversing tumor-induced immunosuppression but may trigger systemic inflammatory responses, increasing neutrophil and platelet counts and fluctuating lymphocyte levels—all contributing to SII score changes. Chemotherapy drugs kill cancer cells but damage healthy cells and cause myelosuppression, altering white blood cell and platelet counts to disrupt SII score balance. Additionally, neoadjuvant therapy-induced increases in inflammatory markers (CRP, IL-6) may correlate with SII score changes, further influencing anastomotic healing.

Although this study did not directly analyze the impact of specific neoadjuvant regimens or cycles on SII scores, the findings confirm that postoperative SII score elevation (especially on day 3) is closely associated with leakage. Clinicians should closely monitor dynamic SII changes in patients after neoadjuvant chemoradiotherapy. Significant SII elevation may indicate severe immune-inflammatory imbalance, requiring proactive perioperative interventions such as optimizing treatment protocols to reduce inflammation and enhancing immunomodulation to lower leakage risk.

### 4.5 Limitations of this study

This study has certain limitations: First, while it is a two-center retrospective study, selection bias may still exist, and the sample size (247 cases) may limit the extrapolation of results. Future multi-center prospective studies with expanded sample sizes could yield more reliable conclusions. Second, although this study analyzed SII scores at four time points (preoperative, postoperative day 1, 3, 7), it lacked continuous monitoring between these time points (e.g., postoperative day 2, 4-6), which may hinder comprehensive understanding of SII’s dynamic correlation with leakage timing. Third, despite comparing SII scores across anastomotic leakage grades (I-IV), the difference was not statistically significant (P=0.57), possibly due to the small number of grade IV cases (3 cases), limiting analysis of the correlation between SII and leakage severity. Fourth, this study did not include factors such as surgical approach, operation duration, and postoperative nutritional status, which may influence anastomotic leakage and compromise model comprehensiveness.

### 4.6 Future research direction

Based on the results and limitations, future research can focus on: First, conducting prospective studies to continuously monitor dynamic SII changes in patients undergoing neoadjuvant chemoradiotherapy, refining the optimal predictive time point and cutoff value to enhance clinical applicability. Second, exploring the impact of different neoadjuvant regimens (immune checkpoint inhibitor combinations, chemotherapy cycles) on SII scores and leakage risk to guide personalized treatment selection. Third, combining SII scores with other inflammatory indicators (CRP, PCT), nutritional indicators (albumin), and clinical factors to construct a more comprehensive risk prediction model for improved early identification of high-risk patients. Fourth, investigating pharmaceutical (anti-inflammatory, immunomodulatory drugs) and non-pharmaceutical (nutritional support) strategies to regulate SII scores and reduce leakage incidence, providing new therapeutic targets.

In conclusion, this study confirms that postoperative day 3 SII score elevation, advanced age, and hypertension are independent risk factors for postoperative anastomotic leakage in esophageal cancer patients after neoadjuvant chemoradiotherapy. Clinicians should prioritize dynamic SII monitoring and management of these risk factors to identify high-risk patients early and implement targeted interventions, thereby reducing leakage incidence and improving patient outcomes.

## Data Availability

Data cannot be shared publicly because of [patients' private data]. Data are available from the Ethics Committees of the two centers (contact via tel: +860535-4770669) for researchers who meet the criteria for access to confidential data.

## Acknowledgments

The authors thank the patients and their families for participating in this study. We greatly thank Dr Jiangbo-Lin, for providing his kind comments for this study.

